# Managing occupational dermatitis during pandemics using virucidal emollient Dermol 500

**DOI:** 10.1101/2021.02.12.21251419

**Authors:** Christine T. Major-Styles, Michael Vanden Oever, Jonathan Brown, Giulia Gallo, Sweta Rai, Sarah Walsh, Finola M. Ryan, Dalan Bailey, Wendy S. Barclay, Rachel S. Edgar

## Abstract

**Background:** Work-related contact dermatitis is a significant problem for healthcare workers. Repeated exposure to water, soap and alcohol-based gels can cause dry skin, irritation and contact dermatitis. Soap substitutes such as Dermol 500 are commonly prescribed to prevent and treat dermatitis. However, hand hygiene recommendations during the COVID-19 pandemic advised against using soap substitutes in clinical settings as their virucidal capacity had not been established.

**Aims:** To investigate whether Dermol 500 lotion exhibits virucidal activity and is a suitable soap substitute for use during viral disease outbreaks. This study aims to inform best practice in occupational medicine and help combat the increased incidence of work-related contact dermatitis in clinical settings.

**Methods:** The virucidal activity of Dermol 500 lotion and its antimicrobial constituents benzalkonium chloride (BAK) and chlorhexidine dihydrochloride (CD) was assessed against influenza A virus (IAV), SARS-CoV-2 and pseudotyped viruses expressing glycoproteins from 2026 outbreak strains of Ebolavirus and Hantavirus. Viruses or pseudoviruses were incubated with treatment prior to quenching serial dilution and infectivity measurement by plaque assay, tissue culture infectious dose 50 (TCID50) or bioluminescent reporter activity.

**Results:** Dermol 500 exhibits comparable virucidal activity to alcohol-based hand sanitisers against IAV and SARS-CoV-2. Furthermore, the Dermol 500 components BAK and CD exhibit independent and synergistic virucidal activity against IAV, SARS-CoV-2 and pseudoviruses.

**Conclusions:** The virucidal activity of Dermol 500 lotion and its active antimicrobials BAK and CD indicates its suitability as a soap substitute to treat and prevent contact dermatitis in front line workers during viral disease outbreaks.

**Teaser text:** Occupational dermatitis soared during the COVID-19 pandemic, but the standard treatment – the antimicrobial emollient and soap substitute Dermol 500 – was not recommended as its antiviral activity was unknown. Here, we address this evidence gap: Dermol 500 lotion is an effective virucide, and its constituents benzalkonium chloride (BAK) and chlorhexidine dihydrochloride (CD) effectively neutralise SARS-CoV-2, influenza A virus and pseudotyped 2026 strains of Ebolavirus and Hantavirus at concentrations 10-fold lower than the active formulation.

## Introduction

Hand hygiene is imperative in healthcare settings and hand washing with soap or use of alcohol-based hand sanitisers are advised worldwide to assist infection control(1). However, such hand hygiene measures and use of personal protective equipment (PPE) such as gloves and masks can precipitate irritant contact dermatitis(2). This skin condition is characterised by damage to the stratum corneum layer of the epidermis, resulting in cracked, dry and itchy skin with impaired barrier function, which advances to severe irritation and inflammation(3). Following the development of dermatitis there is an elevated risk of bacterial colonisation and subsequent transmission to other healthcare workers and/or patients.

Work-related contact dermatitis in healthcare professionals was exacerbated during the COVID-19 pandemic. 84.6% of surveyed Wuhan healthcare workers reported adverse skin reactions attributed to increased hand-washing frequency(4), with 97% front-line heath care workers in China reporting skin damage related to enhanced infection control measures(5). We observed a sharp increase in NHS staff presenting with irritant hand dermatitis from the onset of the COVID-19 pandemic (Figure 1)(6). Between 26^th^ March – 6^th^ May 2020, 468 staff from King’s College Hospital NHS Foundation Trust were diagnosed with irritant hand dermatitis upon presenting to a staff dermatology clinic. For comparison, only 8 staff had this diagnosis recorded over the same period in 2019 at the occupational health department of the hospital. A 9.5-fold increase in irritant contact dermatitis diagnoses was observed overall in 2020 compared to 2019 (Figure 1C), with significantly increased occupational health walk-in cases and diagnoses following management referral (Figures 1A,B).

**Figure 1.**
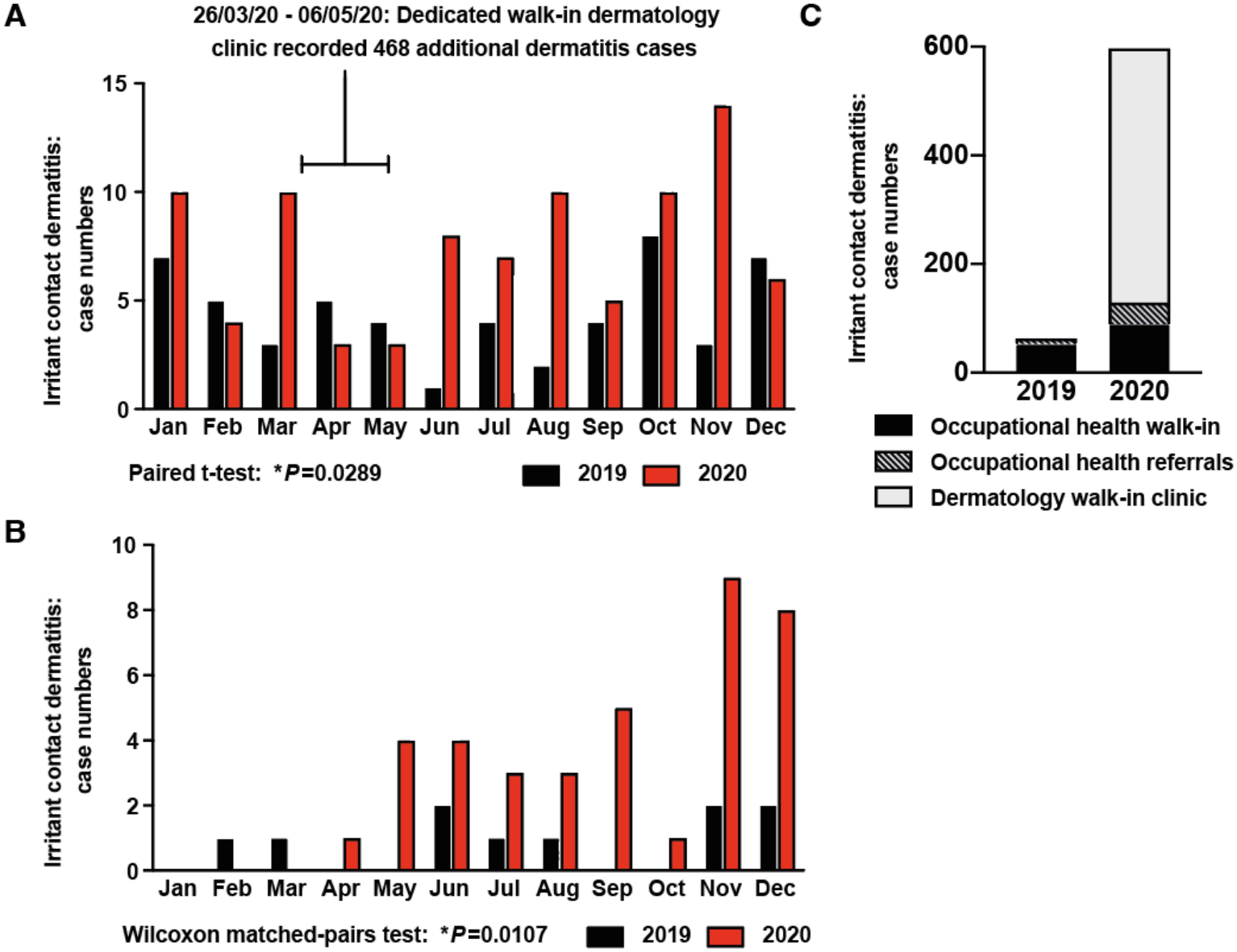
Comparative analysis of staff diagnosed with irritant contact dermatitis at King’s College Hospital NHS Foundation Trust during 2019 and 2020. Irritant contact dermatitis following (**A**) walk-in presentation to occupational health, (**B**) management referral to occupational health, and (**C**) presentation at a dedicated ‘pop up’ staff dermatology clinic established by Drs. Rai and Walsh between 26^th^ March – 6^th^ May 2020.

Although dermatitis is a prominent issue for healthcare workers, it is manageable with appropriate interventions(3). Under UK health and safety law, legislation details that employers have a duty of care and statutory legal obligation to employees to minimise and avoid health problems caused by work, including development of skin conditions such as work-related dermatitis(7). ‘The Control of Substances Hazardous to Health Regulations, (COSHH) 2002, (as amended)’ requires employers to identify substances that may be hazardous to health, assess risks and implement adequate control measures to control exposure and protect health(8). Indeed, if the contact dermatitis is considered solely caused by work, it is reportable under the ‘Reporting of Injuries, Diseases and Dangerous Occurrences Regulations (RIDDOR) 2013’(9, 10). Prior to the COVID-19 pandemic, the use of soap substitutes and emollients was advised to treat work-related contact dermatitis in healthcare workers by the Royal College of Physicians and other medical bodies(2), meeting the COSHH requirement to eliminate exposure to the hazardous substance and substitute with an alternative. Notably, Dermol 500 lotion is commonly prescribed as an antimicrobial emollient and soap substitute to aid skin conditions such as dermatitis(11). However, due to a lack of evidence regarding whether Dermol 500 can inactivate virus particles, its use in clinical settings was discouraged during the COVID-19 pandemic(12).

Dermol 500 contains the antimicrobial components benzalkonium chloride (BAK) and chlorhexidine dihydrochloride (CD), which have well-established antimicrobial activity against *Chlamydia, Pseudomonas, Streptococcus* and *Staphylococcus* bacterial species, amongst others (11, 13-17). BAK alone exhibits virucidal activity against various enveloped viruses, including Canine coronavirus (CCV), herpes simplex virus 1 (HSV1), human immunodeficiency virus (HIV) cytomegalovirus (CMV) and respiratory syncytial virus (RSV)(15, 18, 19). In addition, surface disinfectants where BAK is the principle active component inactivate SARS-CoV-1(20). The parental compound of CD (chlorhexidine) has documented virucidal activity, with alternative salt form derivatives chlorhexidine gluconate and chlorhexidine acetate inactivating HIV and HSV-1(19, 21).

Here, we directly investigate the virucidal activity of Dermol 500 emollient lotion against influenza A virus (IAV) and SARS-CoV-2 in comparison to alcohol-based hand sanitisers, and the efficacy of constituents BAK and CD at the concentrations found within this formulation. Epidemics continually emerge and we evaluated the activity of BAK and CD to neutralise pseudoviruses carrying glycoproteins from the Bundibugyo ebolavirus (BDBV) and Andes hantavirus (ANDV) strains responsible for the recent deadly outbreaks seen in the Democratic Republic of Congo and the MV Hondius cruise ship, respectively (22, 23).

## Methods

### Cell culture

Madin-Darby Canine Kidney (MDCK) cells, VeroE6, 293T-HEK and Huh7 cells were routinely cultured at 37°C with 5% CO2 in Dulbecco’s modified Eagle’s Medium (DMEM; Gibco-Life Technologies) supplemented with 10% fetal calf serum (FCS), 1% penicillin/streptomycin either Glutamax (MDCK) or NEAA (Vero E6) (Gibco-Life Technologies).

### Influenza A virus (IAV) infectivity assay

The influenza A virus used in this study was strain A/PR8/8/34 (H1N1), henceforth referred to as IAV. IAV was propagated in confluent MDCK cells with TPCK-treated trypsin (1 mg/ml; Worthingtons Bioscience) and serum-free DMEM containing 1% penicillin/streptomycin and 1% Glutamax (SFM). MDCK cells were seeded in 6 well plates 24 - 72h prior to IAV infectivity assays. Due to their viscosity, Dermol 500 lotion and commercially available alcohol-based sanitisers were diluted 1:1 in SFM to enable accurate pipetting. IAV was incubated with the specified concentrations of Dermol 500, alcohol-based sanitisers or SFM alone. The alcohol-based hand sanitisers assessed were: 1) Cuticura Original Hand Sanitiser, 2) Lifebuoy Hand Hygiene Sanitiser 3) Hygiene Vision Europe Ltd Hand Hygiene Sanitiser. Alternatively, IAV was incubated with BAK and CD at the concentrations described (Sigma Aldrich; CD suspended in DMSO), culture medium alone or DMSO vehicle control (Figure S1).

Virus was incubated with the treatment for different durations at either room temperature (RT) or 32°C (average skin temperature). Following incubation, the virus/treatment samples were serially diluted in SFM to quench the virucidal activity and infectious IAV determined by standard plaque assay. Briefly, confluent MDCK cells were incubated with IAV dilutions for 1h at 37°C. Virus was then aspirated and cells overlaid with SFM containing 0.8% Avicell (FMC BioPolymer), 0.14% BSA and 1 mg/mL TPCK-treated trypsin (Worthingtons Bioscience). After 3 days, infected cells were fixed in phosphate buffered saline (PBS) with 10% formalin, stained with 0.1% toulidine blue and viral plaque forming units (PFU) assessed. The limit of detection for Dermol 500 and alcohol-based sanitisers was 10^1^ PFU /mL. Due to adverse effects of BAK and CD on cultured in vitro cell lines the limit of detection for BAK and CD treatments between 0.05 - 0.1% was 10^2^ PFU/mL without filtration. Where noted, IAV was filtered to remove BAK/CD using 15 mLlAmicon Ultra-15 centrifugal filter with 50,000 mw cut off (UFC905008, Merck) and centrifugation (4000 × g, 10 min). Filtered virus was resuspended in SFM and infectivity assessed as above.

### SARS-CoV-2 TCID50 infectivity assay

The SARS-CoV-2 strain used in this study was SARS-CoV-2/England/IC19, henceforth referred to as SARS-CoV-2(24). Vero E6 cells were seeded into 96-well plates in supplemented DMEM 24h prior to infectivity assays. SARS-CoV-2 was incubated with either Dermol 500 or alcohol-based sanitiser diluted as for IAV above. Alternatively, BAK and CD diluted to 0.2% in assay diluent (DMEM, 0.3% BSA, 1X NEAA, 1X penicillin/streptomycin) were mixed with an equal volume of SARS-CoV-2 to a final concentration of 0.1% BAK and 0.1% CD. As a control, SARS-CoV-2 was incubated with assay diluent in the absence of Dermol 500, alcohol-based sanitisers or CD/BAK. After the stated incubation period, samples were added to confluent Vero E6 cells and log_10_/half-log_10_ dilution series immediately performed to quench virucidal activity (4 technical replicates/sample). Plates were incubated for 5 days at 37°C before adding crystal violet stain (0.1% w/v) to live cells. Wells were scored for either an intact, stained cell sheet or the absence of cells due to virus-induced cytopathic effect. For each condition, the Spearman-Karber method was used to calculate the 50% tissue culture infectious dose (TCID50) of the virus.

### Pseudovirus infectivity assay

Lentivirus-based pseudovirus containing Andes hantavirus (ANDV) glycoproteins from the 2026 outbreak strain were generated as described previously(25). Briefly, 293T-HEK cells were co-infected with plasmids encoding ANDV envelope glycoprotein, firefly luciferase reporter and pCAGGs-GAG-POL and pseudovirus harvested 48-72h post transfection. rVSV pseudovirus carrying glycoproteins from the 2026 outbreak strain of Bundibugyo ebolavirus (BDBV) were purchased from VirionX LTD. Pseudoviruses were incubated with 0.01% BAK and 0.01% CD, or in combination, for 0.5 minutes at RT before dilution in SFM to quench virucidal activity (Figure S4). Treated pseudoviruses were placed in triplicate onto confluent 293T-HEK (BDBV) or Huh7 (ANDV) cells and incubated for 48 hours before luciferase activity assessed using a Bright-Glo™ kit (Promega) as per manufacturer’s instructions and read on a CLARIOstar^*plus*^ plate reader (BMG Labtech). Triplicate mock infected cells were included on each analysed plate to control for background luminescence.

### Statistical analysis

All data was analysed using GraphPad Prism and presented as mean ± standard deviation (mean±SD). For each treatment condition at least 2 independent biological replicates were performed. Statistical tests for assays were performed on log_10_-transformed data (Y=log_10_[Y+1]).

### Ethical approval

Ethical approval was not required for this study.

## Results

### Dermol 500 lotion exhibits virucidal activity against IAV and SARS-CoV-2 comparable to alcohol-based sanitisers

To assess whether Dermol 500 lotion exhibits virucidal activity, we examined the capacity of IAV and SARS-CoV-2 to infect cells following incubation with this soap substitute (Figure 2). Dermol 500 lotion was diluted in cell culture medium to allow for accurate pipetting, therefore could only be assessed at 25% of its original concentration. Even at this dilution, we observed significant reduction in the amount of infectious IAV after 30 min incubation at skin temperature 32°C (~100-fold reduction in plaque-forming units; Figure 2A), and a similar decrease in SARS-CoV-2 infectivity after only 5 min (Figure 2B). In parallel, we assessed the virucidal activity of three commercially available alcohol-based hand sanitisers. Dermol 500 lotion performed equally or better than these alcohol-based sanitisers at decreasing viral infectivity. The virucidal effect of Dermol 500 was concentration-dependent, showing a smaller significant reduction in IAV infectivity at 12.5% dilution (Figure S2), indicating that efficacy against viruses should be increased when used at its intended concentration.

**Figure 2.**
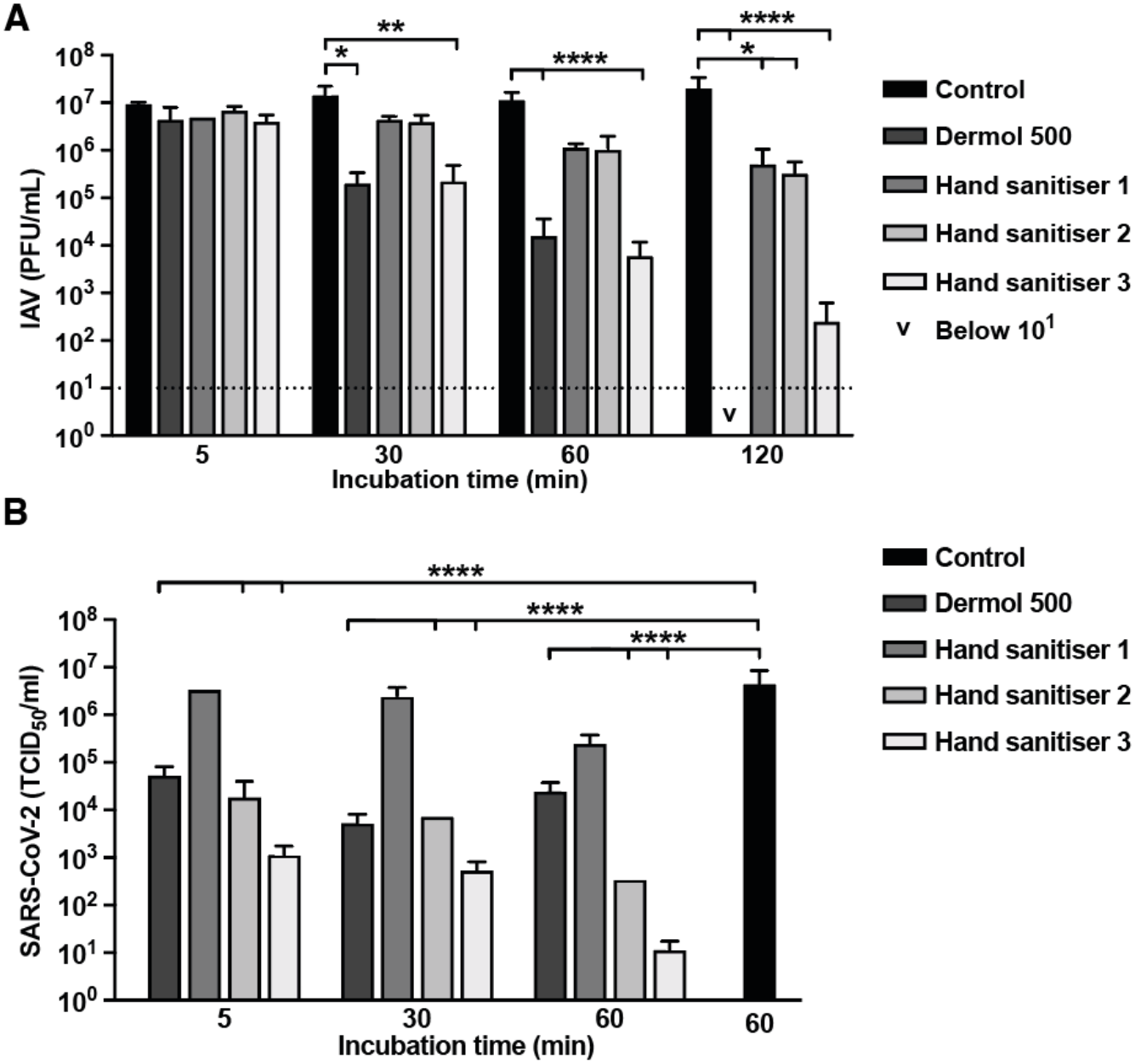
Dermol 500 lotion exhibits virucidal activity against influenza A virus (IAV) and SARS-CoV-2. IAV (**A**) or SARS-CoV-2 (**B**) were incubated with culture medium, alcohol-based hand sanitisers or Dermol 500 lotion at 25% final concentration for 5 - 120 minutes at 32°C. Virus infectivity was assessed by plaque assay for IAV or TCID50 for SARS-CoV-2 (N=2, n=4; mean±SD). For IAV, 2-way ANOVA [treatment × time]: treatment ****P<0.0001, time P>0.05. For SARS-CoV-2, 2-way ANOVA [treatment × time]: treatment ****P<0.0001, time **P<0.0032. Multiple comparisons versus control: *P<0.05, **P<0.01, ***P<0.001, ****P<0.0001. v = below 10^1^ detection limit.

### Dermol 500 antimicrobial constituents benzalkonium chloride (BAK) and chlorhexidine dihydrochloride (CD) rapidly inactivate IAV and SARS-CoV-2

BAK and CD are the active antimicrobial agents in Dermol 500 lotion, each present at 0.1% (w/w)(11). We tested their ability to inactivate IAV and SARS-CoV-2 in combination at this concentration, to determine whether they contribute to the observed virucidal activity of this soap substitute (Figure 3). Infectious IAV decreased 100,000-fold following a 0.5 min incubation with 0.1% BAK/0.1% CD (w/v), and after 3 min we did not detect any viral infectivity (Figure 3A). No SARS-CoV-2-induced cytopathic effect was observed for samples incubated with 0.1% BAK/0.1% CD (w/v) across all incubation time frames (10^2^ TCID50/mL detection limit), a >10,000-fold reduction in virus infectivity within 0.5 min. We conclude that BAK and CD together are extremely potent virucidal agents against influenza and coronaviruses at the concentrations found within Dermol 500 lotion.

**Figure 3.**
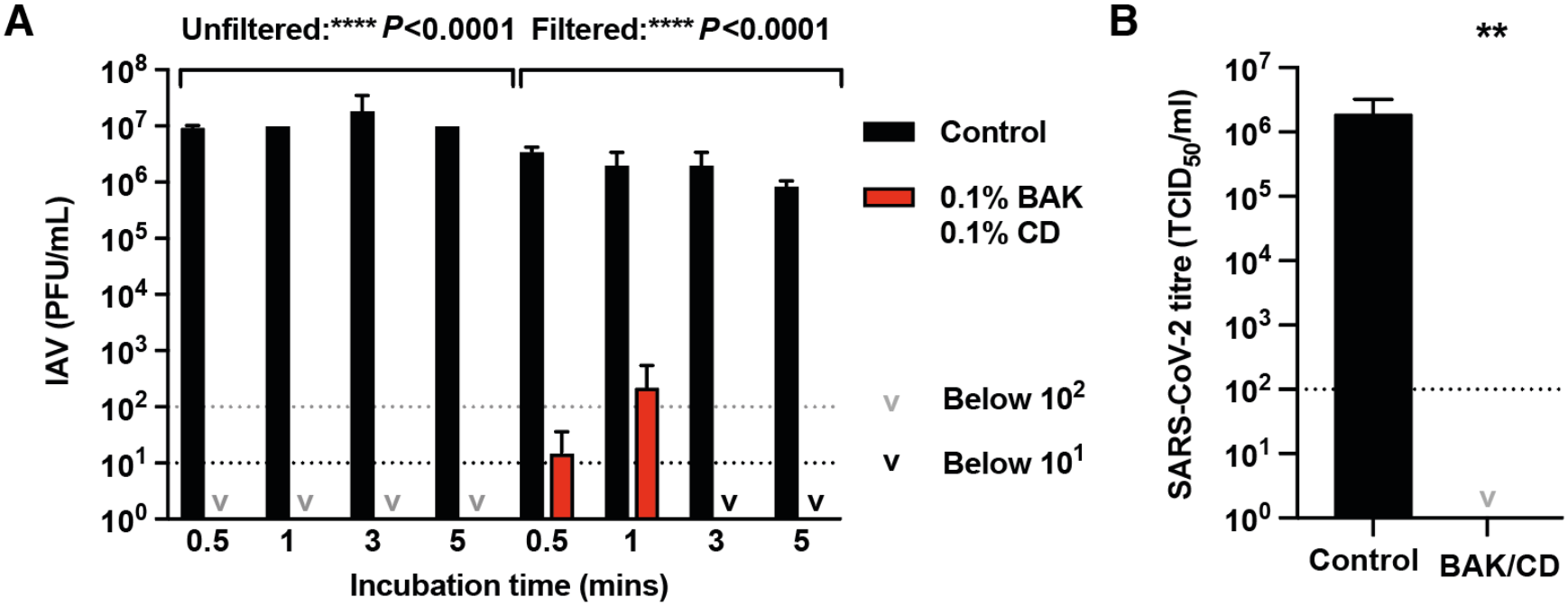
Benzalkonium chloride (BAK) and chlorohexidine dihydrochloride (CD) exhibit rapid virucidal activity against IAV and SARS-CoV-2 at 0.1% concentration. (**A**) IAV was incubated at 32°C with 0.1% BAK + 0.1% CD, or medium only control, for 1-min. IAV infectivity was assessed by plaque assay directly (unfiltered; 10^2^ PFU/mL detection limit; N=2; mean±SD), or parallel samples were subject to filtration to remove BAK/CD prior to plaque assay (filtered; 10^1^ PFU/mL detection limit; N=2; mean±SD). 2-way ANOVA [treatment × time]: unfiltered treatment ****P<0.0001, time P>0.05; filtered treatment ****P<0.0001, time P>0.05. (**B**) SARS-CoV-2 was incubated at RT with 0.1% BAK + 0.1% CD for either 0.5 min (N=2), 10 min (N=2) or 30 min (N=4), or assay diluent control for 30 min (N=4)(mean±SD). Virus infectivity was assessed by TCID50 assay. No cytopathic effect was observed with BAK/CD treated virus across all incubation time frames (10^2^ TCID50/mL detection limit). 2-tailed Mann-Whitney test: **P=0.002.

### BAK and CD have different virucidal efficacies and act synergistically to disrupt viral infectivity

We assessed the virucidal activity of decreasing concentrations of BAK and CD, in combination and independently to further characterise their efficacy (Figure 4). Within 0.5 min, we observed a significant decrease in IAV infectivity with the lowest concentration of BAK tested (0.0125% w/v), indicating that amounts could be substantially reduced when formulating virucidal soap substitutes and sanitisers. CD alone did not decrease infectious virus over the 0.5 min time frame but does show synergistic interaction with BAK by enhancing its efficacy against IAV in a time- and temperature-dependent manner (Figure 4; Figure S3). At 32°C (skin temperature), CD exerts an independent virucidal effect at higher concentrations after 5 min and the lowest CD concentration tested reduced IAV infectivity by >100-fold within 30 min. 0.0125% BAK/0.1% CD (w/v) was the most efficient combination tested, with infectious IAV below detectable levels in 0.5 min (Figure S3).

**Figure 4.**
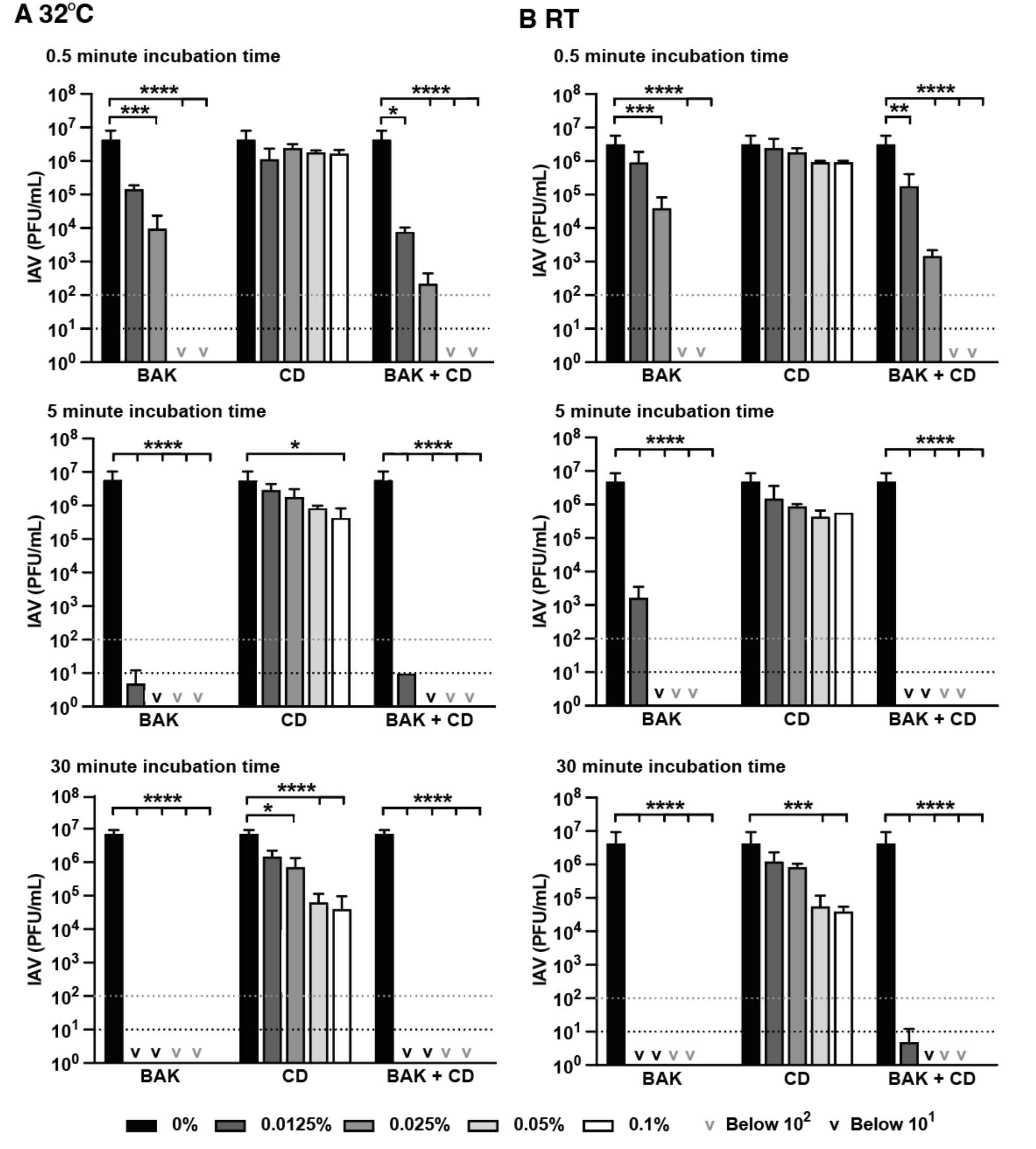
Synergistic virucidal activity of benzalkonium chloride (BAK) and chlorohexidine dihydrochloride (CD) at concentrations less than 0.1%. IAV was incubated at 32°C (**A**) or RT (**B**) with decreasing concentrations of BAK, CD or in combination for 0.5-30 minutes. Infectivity was assessed by plaque assay (N≥2; mean±SD). 32°C and RT 2-way ANOVA [% drug × time]: drug ****P<0.0001, % ****P<0.0001. Multiple comparisons versus 0%: *P<0.05, **P<0.01, ***P<0.001, ****P<0.0001.

Outbreaks of emerging viral strains with epidemic and pandemic potential are a continuous threat to public health. Therefore, we explored the broader context of BAK and CD activity against disease causing viruses, utilizing pseudovirus systems engineered to carry glycoproteins from the 2026 viral outbreaks of ANDV hantavirus and BDBV ebolavirus (22, 23). Given the robust virucidal activity found in our prior experiments and the dilution constraints of quenching virucidal activity prior to cell-based bioluminescence assays for pseudovirus infectivity (Figure S4), we treated pseudoviruses with 0.01% BAK/CD (w/v) final concentration independently and in combination for 0.5 minutes before quenching and infection of susceptible cells. Consistent with robust virucidal activity against IAV and SARS-CoV-2 (Figure 2-4), 0.01% BAK and CD in combination significantly reduced infectivity of hantavirus and ebolavirus pseudoviruses following 0.5 min incubation (Figure 5).

**Figure 5.**
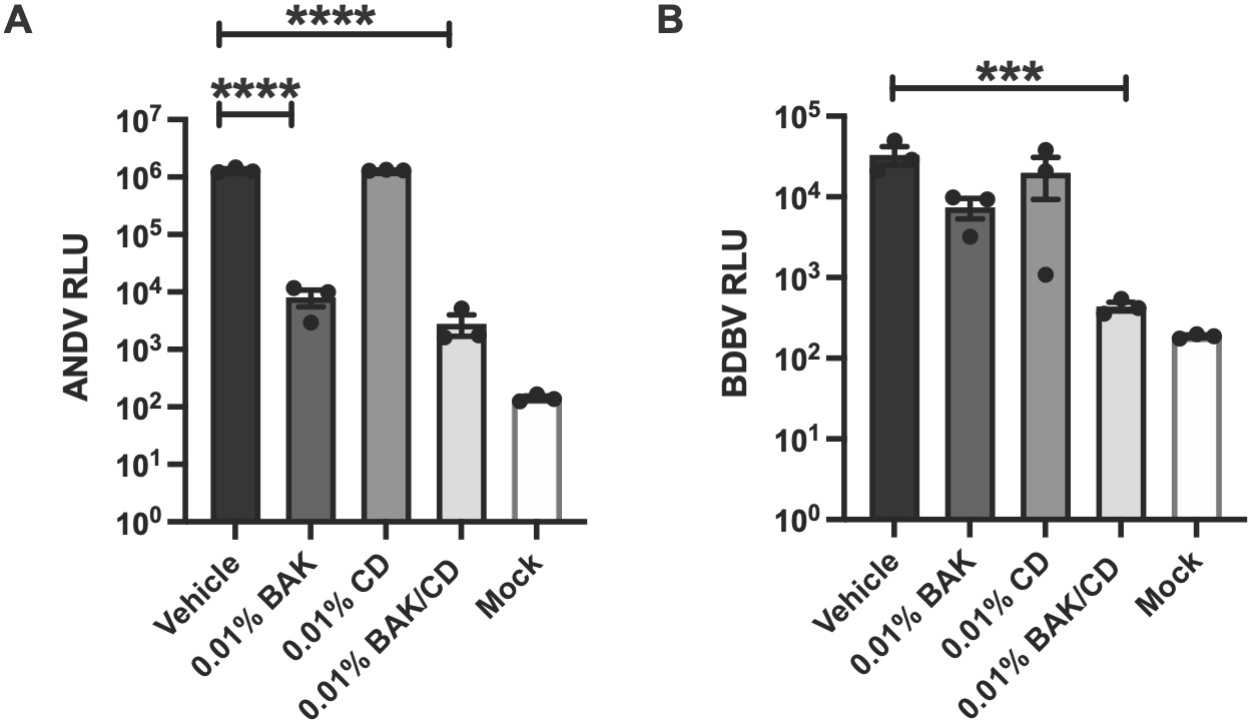
Benzalkonium chloride (BAK) and chlorohexidine dihydrochloride (CD) exhibit broad-spectrum virucidal activity. Pseudoviruses expressing glycoproteins from ANDV hantavirus (**A**) or BVBD ebolavirus (**B**) were incubated with 0.01% BAK, 0.01% CD, 0.01% BAK + 0.01% CD, or vehicle control, for 0.5 minutes at RT. Infectivity was assessed via bioluminescence (N=3, n=3; mean±SD). White bars = mock infected. 1-way ANOVA [BAK/CD], multiple comparisons versus vehicle control: ***P<0.001, ****P<0.0001. RLU; relative luciferase units.

## Discussion

This study clearly demonstrates Dermol 500 emollient lotion, and its antimicrobial components BAK and CD, exhibit robust virucidal activity against IAV and SARS-CoV-2, and significantly reduce the infectivity of pseudoviruses carrying glycoproteins from 2026 outbreak strains of ebolavirus and hantavirus. These findings support the ongoing use of Dermol 500 as a soap substitute to treat, manage and prevent occupational contact dermatitis during viral epidemics.

The virucidal activity of Dermol 500 lotion is mediated by the synergistic activity of BAK and CD. BAK is known to disrupt the physical integrity and biochemical function of bacterial lipid membranes (13, 15, 26-29) so could target the viral envelope. This compound is also effective against several non-enveloped viruses, suggesting an effect on protein structure(15, 19, 30). This virucidal activity requires longer incubation and higher concentrations than for enveloped viruses, in line with inefficient inactivation of non-enveloped viruses by alcohol-based and other disinfectants(1, 31). The virucidal mechanism of action of CD is not established, but chlorhexidine antimicrobial activity is associated with denaturation of membrane bound proteins so may disrupt viral glycoprotein conformation, particularly in the presence of BAK(13, 32). Delineating how BAK and CD reduce virus particle infectivity is beyond the scope of this paper, but their synergistic activity reported here indicates they have complementary mechanisms of action. Additionally, BAK is an effective virucide against enveloped viruses and pseudoviruses at 10-fold lower concentrations than found in Dermol 500 lotion, useful for future development of soap substitutes and other topical infection control products. Interestingly, BAK is a component of many commercially available nasal sprays at a highly effective concentration of 0.1% w/v, listed as a preservative rather than an active ingredient.

Hand hygiene and infection control measures involve repeated exposure to water and use of soap, alcohol hand gels and other detergents. Exposure to these irritants and PPE such as masks and gloves elevates the risk of work-related skin disease, with irritant contact dermatitis the most common occupational inflammatory skin disease in Europe(33). During the COVID-19 pandemic, these infection control measures were widely adopted across the globe and increased occurrence of work-related contact dermatitis likely extended beyond clinical settings into commercial employment industries, particularly other key workers. Occupational dermatitis is a reportable disease under UK health and safety legislation(7, 9), and employers are legally obliged to minimise exposure to harmful substances and reduce the risk of health problems including skin conditions. By establishing the virucidal efficacy of a common treatment option, this study enables future field studies to address the utility of emollient soap substitutes like Dermol 500 in curbing the large increases in health worker skin conditions observed during viral pandemics (Figure 1;(4, 5)). Ultimately, a hand hygiene alternative that combats both viruses and skin irritation would decrease staff sickness absences and restriction from clinical work due to skin disease, whilst enhancing staff wellbeing and infection control compliance for the safe and timely delivery of clinical care.

## Key learning points

### What is already known about this subject

- Hand hygiene requirements in clinical settings increase the risk of irritant and work-related contact dermatitis.
- Dermol 500 is widely prescribed as an emollient and soap substitute for dermatitis management, but its continued use during the COVID-19 pandemic was not recommended as its ability to neutralise SARS-CoV-2 and other viruses was unknown.
- There is a requirement for dermatitis treatments that are compatible with enhanced infection control measures during viral outbreaks.

### What this study adds

- Occupational data showing a significant increase in irritant dermatitis diagnoses in healthcare workers at a large London hospital during 2020 compared to 2019.
- The common dermatitis treatment Dermol 500 lotion effectively reduces SARS-CoV-2 and influenza A virus infectivity, with comparable activity to alcohol-based sanitisers.
- Dermol 500 constituents benzalkonium chloride and chlorhexidine dihydrochloride exhibit rapid, synergistic and broad-spectrum virucidal activity against SARS-CoV-2, IAV and pseudotyped 2026 strains of hanta- and ebolaviruses responsible for the 2026 outbreaks.

### What impact this may have on practice or policy

- This study supports continued use of Dermol 500 lotion as a soap substitute during viral outbreaks, enabling the evaluation of its utility in managing work-related dermatitis under these conditions that elevate dermatological risk.
- Developing an integrated approach to hand hygiene and skin irritation management for healthcare workers to reduce sickness absence, improve staff wellbeing and support compliance with infection control measures.

## Data Availability

Data is available on request.

## Acknowledgments

We thank Dr Lucia Batty, Dr Lisa Curran and Mr James Kesson (King’s College Hospital NHS Foundation Trust, London) for their assistance during the preparation of this manuscript, and Dr John O’Neill (MRC Laboratory of Molecular Biology, Cambridge) for helpful discussion.

## Supporting information

**Figure S1.**
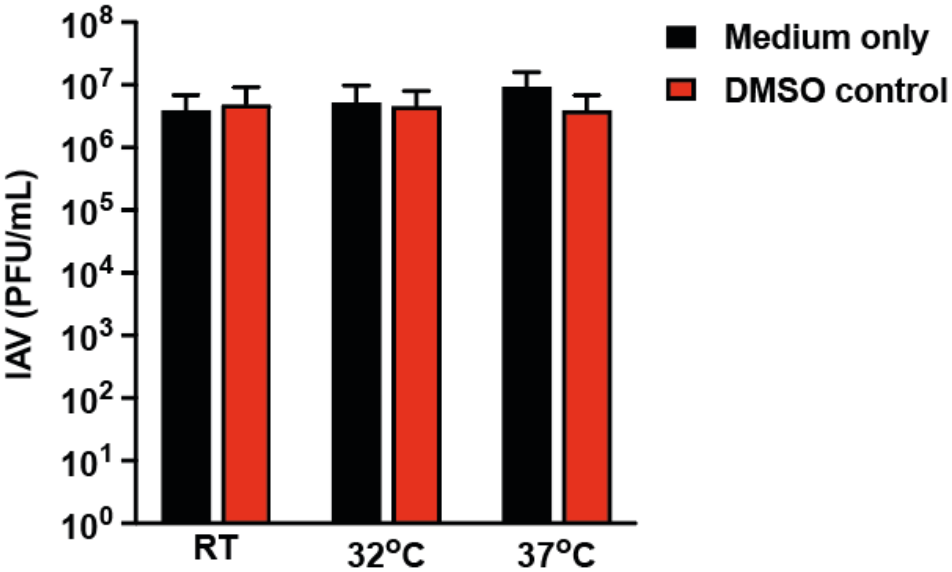
Chlorohexidine dihydrochloride (CD) vehicle DMSO does not exhibit virucidal activity. Influenza A virus (IAV) was incubated with 2% DMSO (vehicle concentration equivalent to 0.1% CD) or medium-only control for 120 minutes at room temperature RT, 32°C or 37°C. Virus infectivity was assessed by plaque assay (n=2, mean±SD). 2-way ANOVA (treatment × temperature): P>0.05.

**Figure S2.**
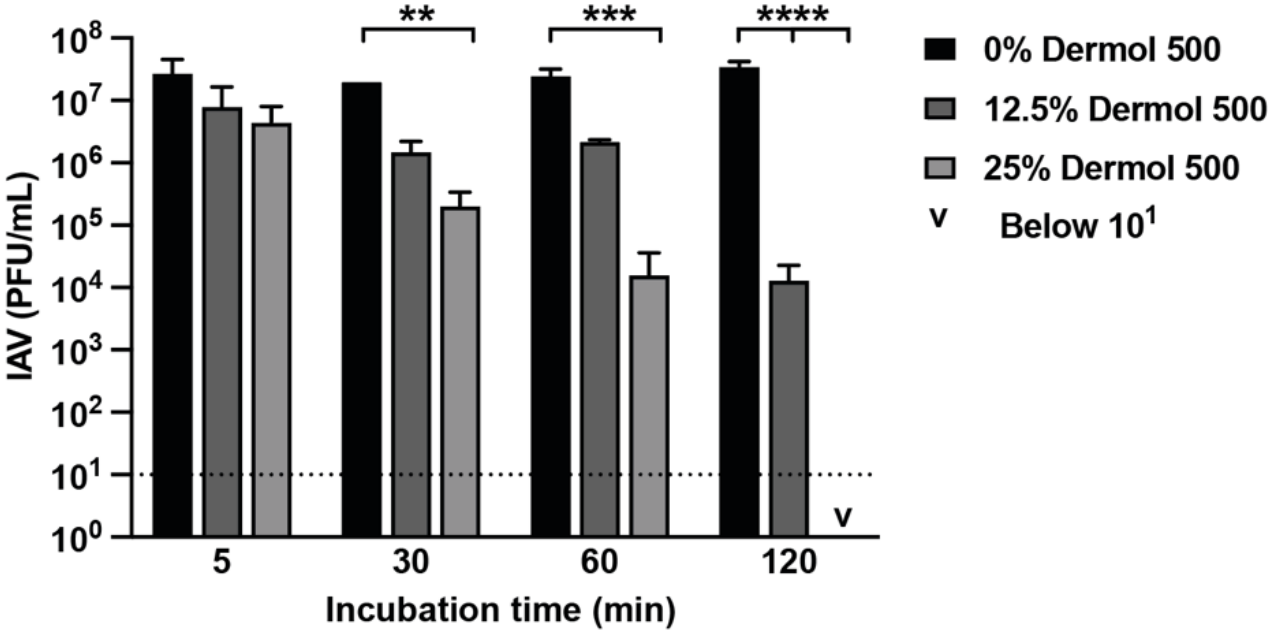
Virucidal activity of Dermol 500 lotion at increasing concentrations. Dermol 500 was diluted in serum-free media (SFM) and incubated with influenza A virus (IAV) at 12.5% or 25% final concentration for 5 – 120 min at 32°C. Virus infectivity was assessed by plaque assay. 2-way ANOVA (time × concentration): concentration **P=0.002, time P>0.05.

**Figure S3.**
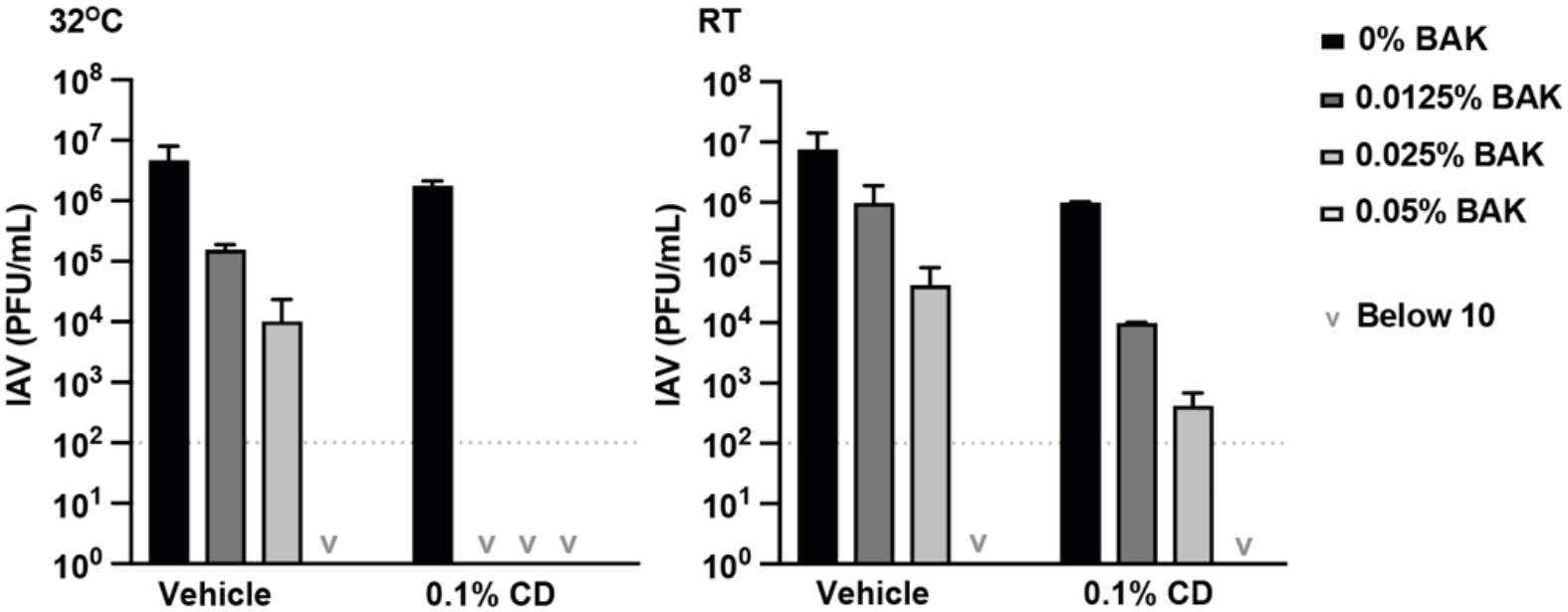
Synergistic virucidal activity of benzalkonium chloride (BAK) and chlorohexidine dihydrochloride (CD). Influenza A virus was incubated for 0.5min at either 32°C or RT with increasing concentrations of BAK (0.0125 – 0.05%), and either DMSO vehicle or 0.1% CD. Virus infectivity was assessed by plaque assay (n=2; mean±SD). 2-way ANOVA [treatment × concentration]: 0.5min treatment P>0.05, concentration **P<0.0018.

**Figure S4.**
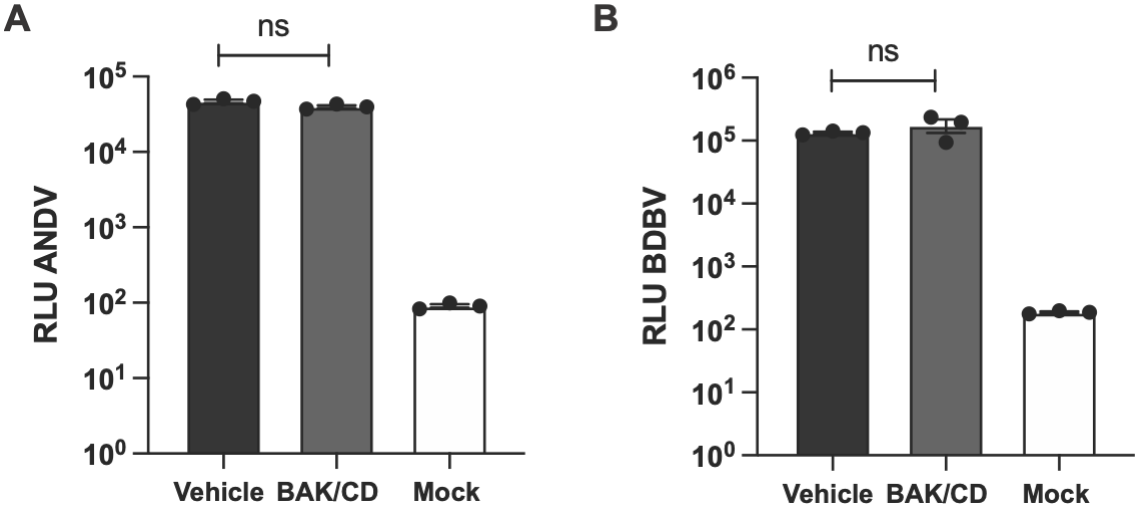
benzalkonium chloride (BAK) and chlorohexidine dihydrochloride (CD) virucidal activity effectively quenched by dilution to 0.0002%. Pseudoviruses expressing glycoproteins from ANDV hantavirus (**A**) or BVBD ebolavirus (**B**) were incubated with 0.00002% BAK and CD in combination, or vehicle control, for 48h assay time frame. Infectivity was assessed via bioluminescence (N=3, n=3; mean±SD). White bars = mock infected. 1-way ANOVA [BAK/CD], multiple comparisons versus vehicle control. ns; non-significant. RLU; relative luciferase units.

